# Folate (*MTHFR* C677T and *MTRR* A66G) gene polymorphisms and risk of prostate cancer: a case-control study with an updated meta-analysis

**DOI:** 10.1101/2021.01.06.21249185

**Authors:** Upendra Yadav, Pradeep Kumar, Shailendra Dwivedi, Bhupendra Pal Singh, Vandana Rai

## Abstract

**Introduction:** Methylenetetrahydrofolate reductase (MTHFR) and methionine synthase reductase (MTRR) are the key enzymes of the folate pathway, which involved in the DNA methylation. DNA methylation may affect the stability and integrity of DNA, that supposed to play a pivotal role in carcinogenesis. So, we aimed to investigate the association of *MTHFR* C677T and *MTRR* A66G gene polymorphisms with susceptibility to prostate cancer in North Indian population. We also performed meta-analyses of published literatures on these polymorphisms to evaluate their association with prostate cancer.

**Methods:** We genotyped *MTHFR* C677T and *MTRR* A66G gene polymorphisms in 147 prostate cancer cases and 147 healthy controls using PCR-RFLP methods. Odds ratios (ORs) with 95% confidence intervals (CIs) were estimated for risk estimation. For meta-analysis different databases were searched and all statistical analysis were performed using Open Meta-Analyst software.

**Results:** The present case control study revealed that the T allele (OR= 1.67; 95% CI: 0.99-2.84, p= 0.05), CT genotype (OR= 1.92; 95% CI: 1.06-3.48, p= 0.02), and dominant (TT+CT) model (OR= 1.85; 95% CI: 1.05-3.30, p= 0.03) of *MTHFR* C677T gene polymorphism and G allele (OR= 1.92; 95% CI: 1.35-2.73, p= 0.0002) of *MTRR* A66G gene polymorphism were significantly associated with prostate cancer susceptibility. Meta-analyses of *MTHFR* C677T and *MTRR* A66G gene polymorphisms showed no significant association between these polymorphisms and prostate cancer risk in overall or in subgroup meta-analysis stratified by ethnicity.

**Conclusion:** *MTHFR* C677T and *MTRR* A66G gene polymorphisms seem to play a significant role in prostate cancer susceptibility in North Indian population, while results of meta-analysis revealed no association between *MTHFR* C677T and *MTRR* A66G gene polymorphisms and prostate cancer susceptibility.

## 1. Introduction

Prostate cancer is a common cancer in men with an estimated 1.2 million new cases and 0.35 million deaths worldwide [1]. The incidence of prostate cancer widely varies in different populations and more common in the developed countries. In India the five year prevalence rate of prostate cancer is 8.1 [1]. Most of the cases of prostate cancer occur in men over the age of 50. Though it is a common cancer but its etiology is poorly understood. Age, ethnicity, and family history of prostate cancer are some of the risk factors. Smoking, occupational chemicals, diet, inflammation, androgens and obesity are considered as secondary risk factors [2].

There are some factors which might be responsible for the development of cancer including DNA hypo-methylation [3], uracil mis-incorporation, and DNA strand break [4]. These all events are linked with the deficiency of the folic acid. The defects in the folic acid pathway leads to the development of cancer, but the exact mechanism is not fully elucidated. Methylenetetrahydrofolate reductase (MTHFR) is an important enzyme of folate pathway that plays a crucial role in DNA synthesis and DNA methylation. Defects in this enzyme may lead to hypo-methylation of DNA that ultimately resulted in the altered genes expression. M*THFR* gene is located on the chromosome 1. A number of polymorphisms are reported in this gene but clinically most important one is *MTHFR* C677T. The other crucial enzyme of the folic acid pathway is methionine synthase reductase (MTRR) which activates the inactive methionine synthase (MTR) enzyme. *MTRR* gene is a housekeeping gene and located at chromosome 5.

The frequency of the *MTHFR* C677T gene polymorphism greatly varies in different population of the world. The frequency of T allele and TT genotype was reported lowest in the African population and highest in the European population [5]. In the Eastern Uttar Pradesh population the frequency of T allele was in the range of 4% to 14% [6-9]. Similarly, the frequency of the *MTRR* A66G gene polymorphism varied from population to population. The lowest frequency of G allele and GG genotype was reported in the South American population and highest in the Asian population [10]. In the Eastern Uttar Pradesh population the frequency of G allele was in the range of 58% to 70% [11-13].

*MTHFR* C677T gene polymorphism is known to be associated with predisposition for different types of cancers like-oral cancer [14], breast cancer [15,16], colorectal cancer [17,18], gastric cancer [19,20], lung cancer [21,22], pancreatic cancer [23,24], and bladder cancer [25,26]. Similarly *MTRR* A66G gene polymorphism is also found to be associated with various types of cancers such as-cervical cancer [27], lung cancer [28,29], meningioma [30], colorectal cancer [31,32], head and neck cancer [33], pancreatic cancer [23], and breast cancer [34].

A number of studies were conducted to check the association of *MTHFR* C677T gene polymorphism with the prostate cancer [35-47]. Very few studies were published, which evaluated *MTRR* A66G polymorphism as risk for prostate cancer [39,40,48-50]. The results of these studies were contradictory for both the selected (*MTHFR* C677T and *MTRR* A66G gene) polymorphisms. Moreover, only a single study Mandal et al. [43] was published from India, which evaluated *MTHFR* C677T polymorphism in prostate cancer patients, and as per our knowledge, from India no study was published so far which evaluated the role of *MTRR* A66G gene polymorphism in the etiology of prostate cancer. So, we designed this study to check the role of *MTHFR* C677T and *MTRR* A66G gene polymorphisms in the North Indian prostate cancer patients and we also performed meta-analyses to check the effect of these polymorphisms on the etiology of prostate cancer.

## 2. Materials and Methods

### 2.1 Genotyping study

#### 2.1.1 Sample collection

A total of 147 prostate cancer cases (age criteria: 40-78 years) were recruited from the outpatient clinic of King George Medical University, Lucknow. In the same time frame, 147 age matched controls from Eastern Uttar Pradesh population were enrolled. Controls were health individuals without any family history and are unrelated to the patients. Informed written consent was obtained from each subjects and the ethical approval was obtained from the Institutional Ethical Committee of King George Medical University, Lucknow and Veer Bahadur Singh Purvanchal University, Jaunpur.

#### 2.1.2 Genomic DNA extraction

3 ml blood sample was collected in EDTA coated vials from both the case and control groups. Genomic DNA was extracted by the method of Bartlett and White [51].

#### 2.1.3 Genotyping

*MTHFR* C677T genotyping was carried out by PCR-RFLP method of Frosst et al. [52], for *MTRR* A66G genotyping the method of Wilson et al. [53] was adopted. Briefly, 100ng of genomic DNA was amplified in a final volume of 15μl with 4pM of each of forward and reverse primers, 250μl of dNTPs mix, 1X *Taq* DNA polymerase buffer and 1U of *Taq* DNA polymerase. PCR program was initial denaturation at 94^0^C for 4 minutes followed by 30 cycles of denaturation at 94^0^C for 1 minute, annealing for 1 minute (at 62^0^C for *MTHFR* C677T and 64^0^C for *MTRR* A66G), extension at 72^0^C for 1 minute and a final extension at 72^0^C for 10 minutes. The amplicons (198bp) were digested with *Hinf*I as the C677T mutation creates a restriction site for it, and resolved in a 2% agarose gel. For A66G amplicons (66bp) digestion were performed with *Nde*I and resolved in a 4% agarose gel.

#### 2.1.4 Statistical analysis

Allele frequencies were calculated by the gene counting method. χ^2^ test was performed to test the Hardy-Weinberg Equilibrium (HWE). Odds ratios (OR) with 95% confidence intervals (CI) were calculated for the comparison of allele and genotype of cases and controls. All statistical analysis was performed by OpenEpi program.

### 2.2 Meta-Analysis

#### 2.2.1 Searched strategy and identification of studies

For meta-analysis PubMed, Science Direct, Springer Link and Google scholar databases were searched for the suitable articles using the combination of the keywords “*MTHFR”*, “methylenetetrahydrofolate reductase”, “C677T”, “*MTRR*”, “methionine synthase reductase”, “A66G” along with “prostate cancer”. The databases were searched up to 31^st^ June, 2020.

#### 2.2.2 Inclusion and exclusion criteria

For a study to include in the meta-analysis, the study should: i) be original; ii) be a case-control study; and iii) be reported *MTHFR* C677T or *MTRR* A66G alleles/genotypes. Studies were excluded if they were: i) reported either only cases or controls, and ii) review, editorial etc.

#### 2.2.3 Data extraction

From all the eligible studies, following information were extracted: family name of the first author, year of publication, country of study, population/ethnic group, number of alleles and/or genotypes.

#### 2.2.4 Statistical analysis

Crude odds ratio (OR) along with the 95% confidence interval (CI) were used to assess the strength of the association between *MTHFR* C677T and *MTRR* A66G gene polymorphisms with the risk of prostate cancer. In the present study, we calculated five genetic models i.e. log additive, homozygote, heterozygote, dominant and recessive models. The ORs were estimated for both the fixed effect [54] and random effect [55] models. The between studies heterogeneity was tested using the Q-statistics and was quantified using the I^2^ statistics [56]. If I^2^ > 50% then random effect model was used otherwise fixed effect model was adopted. We further stratified our results on the basis of ethnicities. For the assessment of publication biases Egger’s test was used [57]. Funnel plot of standard error by log OR and funnel plot of precision by log OR were generated. All the p values were two tailed with a significance level at <0.05. All statistical analyses were performed through Open Meta-analyst program [58].

## 3. Results

For the present case-control study we collected 147 prostate cancer samples (age= 62.72 ± 11.84 years) and the mean prostate specific antigen was 29.61 ± 21.41 ng/ml and same number of age matched controls (age= 62.03 ± 10.76 years).

### 3.1 Association of *MTHFR* C677T and *MTRR* A66G gene polymorphisms with prostate cancer risk

The genotype distribution of *MTHFR* C677T gene polymorphism was in agreement with Hardy-Weinberg equilibrium (HWE) (p= 0.38) but the genotype distribution was deviated from the HWE for the *MTRR* A66G gene polymorphism (p= 00001). For *MTHFR* C677T gene polymorphism, the frequency of C and T allele in cases was 0.86 and 0.14 respectively, while in controls C and T allele frequencies were 0.91 and 0.09 respectively. The CC, CT and TT genotypic frequencies in cases were 0.74, 0.25 and 0.01 respectively, and the frequencies in controls were 0.84, 0.15 and 0.01 respectively. The T allele of *MTHFR* C677T was slightly associated with the prostate cancer (OR= 1.67; 95%CI: 0.99-2.84, p= 0.05). A significant association were also found between CT genotype (OR= 1.92; 95% CI: 1.06-3.48, p= 0.02) and dominant model (OR= 1.85; 95% CI: 1.05-3.30, p= 0.03) and prostate cancer (Table 1).

**Table 1:** Distribution and odds ration with 95% confidence intervals for *MTHFR* C677T and *MTRR* A66G gene polymorphism

|  | Allele/Genotype | Case | Control | OR (95% CI with p-value) |
| --- | --- | --- | --- | --- |
| ***MTHFR* C677T** | CC | 108 | 123 | 1 (Reference) |
|  | CT | 37 | 22 | **1.92 (1.06-3.48), 0.02** |
|  | TT | 2 | 2 | 1.14 (0.12-11.09), 0.89 |
|  | TT+CT | 39 | 24 | **1.85 (1.05-3.30), 0.03** |
|  | C | 253 | 268 | 1 (Reference) |
|  | T | 41 | 26 | **1.67 (0.99-2.84), 0.05** |
| ***MTRR* A66G** | AA | 11 | 9 | 1 (Reference) |
|  | AG | 51 | 96 | 0.43 (0.16-1.14), 0.07 |
|  | GG | 85 | 42 | 1.66 (0.61-4.36), 0.29 |
|  | GG+AG | 136 | 138 | 0.81 (0.31-2.04), 0.64 |
|  | A | 73 | 114 | 1 (Reference) |
|  | G | 221 | 180 | **1.92 (1.35-2.73), 0.0002** |

In *MTRR* A66G gene polymorphism analysis, the frequency of A and G allele was 0.25 and 0.75 respectively in cases while in controls it was 0.39 and 0.61 respectively. The AA, AG and GG genotypic frequencies in cases were 0.07, 0.35 and 0.58 and in control were 0.06, 0.65 and 0.29 respectively. The G allele of *MTRR* A66G gene polymorphism was significantly associated with the prostate cancer (OR= 1.92; 95% CI: 1.35-2.73, p= 0.0002), while no such association was found with any other genetic model (Table 1).

### 3.2 Meta-analysis of *MTHFR* C677T gene polymorphism in prostate cancer

For *MTHFR* C677T meta-analysis we found 25 studies with 12,488 cases and 13,906 controls. [35-49, 59-67, present study]. Out of 25 studies, eight studies were from Asia, 12 were carried out in Caucasians subjects and five studies were of mixed ethnicities. Insignificant association was found in the C677T gene polymorphism in all the genetics models with high heterogeneity (For allele contrast model OR_Tvs.C_= 0.93, 95%CI: 0.85-1.01, p= 0.12, I^2^= 66.96%; for co-dominant model OR_CTvs.CC_= 0.97; 95%CI: 0.86-1.10; p= 0.70; I^2^= 66.77%; for homozygote model OR_TTvs.CC_= 0.80; 95%CI= 0.67-0.97; p= 0.02; I^2^= 57.26%; for dominant model OR_TT+CTvs.CC_= 0.94; 95%CI: 0.84-1.06; p= 0.36; I^2^= 68.6%; and for recessive model OR_CC+CTvs.TT_= 0.83; 95%CI= 0.70-0.98; p= 0.03; I^2^= 53.82%) (Figure 1, Table 2).

**Table 2:**
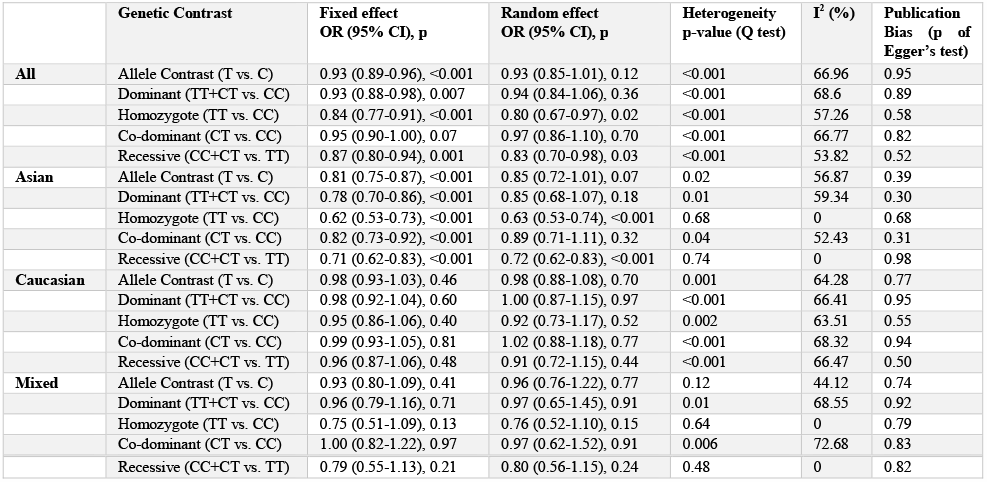
Summary estimates for the odds ratio (OR) of *MTHFR* C677T in various allele/genotype contrasts, the significance level (p value) of heterogeneity test (Q test), and the I^2^ metric

**Figure 1.**
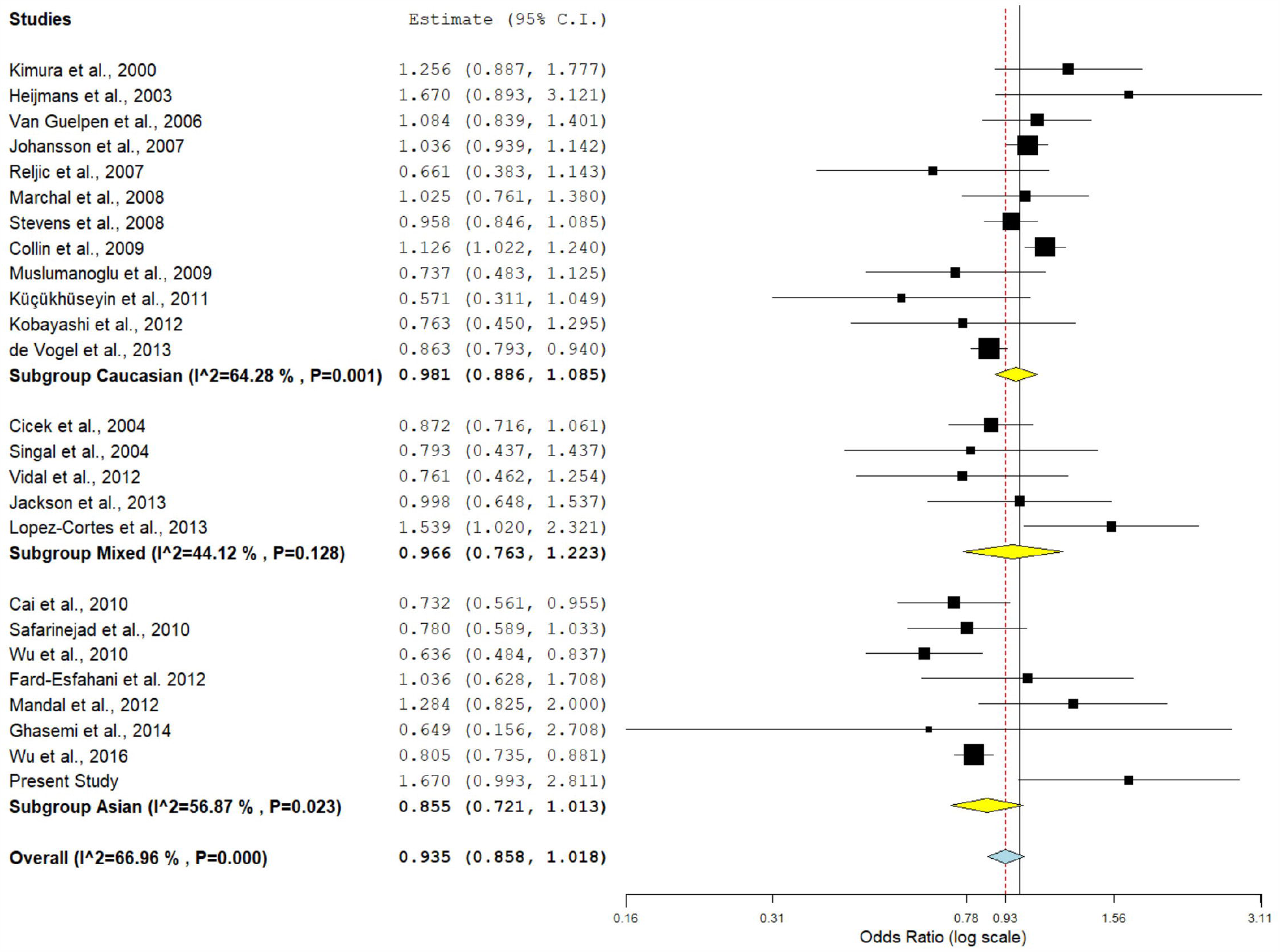
Random effect forest plot of allele contrast model (T vs. C) of *MTHFR* C677T gene polymorphism. Results of individual and summary OR estimates, and 95% CI of each study were shown. Horizontal lines represented 95% CI, and dotted vertical lines represent the value of the summary OR.

In sub-group meta-analyses, no statistically significant association were found in any ethnic subgroup, neither in Asian population (for T vs. C: OR= 0.84; 95% CI=0.72–1.01; p= 0.07; I^2^= 56.87%; for TT + CT vs. CC: OR= 0.85; 95% CI= 0.68-1.07; p= 0.18; I^2^= 59.34%; for TT vs. CC: OR= 0.62; 95% CI= 0.53-0.73; p= <0.001; I^2^= 0%; for CT vs. CC: OR= 0.99; 95% CI=0.71-1.11; p= 0.32; I^2^= 52.43%; and for CT+CC vs. CC: OR= 0.71; 95% CI= 0.62-0.83; p=<0.001; I^2^= 0%) nor in Caucasian population (for T vs. C: OR= 0.98; 95% CI= 0.88-1.08; p= 0.70; I^2^= 64.28%; for TT + CT vs. CC: OR= 1.00; 95% CI= 0.87-1.15; p= 0.97; I^2^= 66.41%; for TT vs. CC: OR= 0.92; 95% CI= 0.73-1.17; p= 0.52; I^2^= 63.51%; for CT vs. CC: OR= 1.02; 95% CI= 0.88-1.18; p= 0.77; I^2^= 68.32%; and for CT+CC vs. CC: OR= 0.91; 95% CI= 0.72-1.15; p=0.44; I^2^= 66.47%) (Table 2).

### 3.3 Meta-analysis of *MTRR* A66G gene polymorphism in prostate cancer

For *MTRR* A66G meta-analysis, total eight studies with 3,631 cases and 5,076 controls [39, 40, 45, 48, 49, 50, 62, present study]. Out of which, three were Asian, three were Caucasian and two were of mixed ethnicities. Insignificant association was found in the A66G gene polymorphism in all the genetics models with high heterogeneity (For allele contrast model OR_Gvs.A_= 1.03; 95%CI: 0.90-1.18; p= 0.59; I^2^= 62.43%; for co-dominant model OR_AGvs.AA_= 0.90; 95%CI: 0.74-1.10; p= 0.32; I^2^= 56.11%; for homozygote model OR_GGvs.AA_= 0.95; 95%CI= 0.83-1.08; p= 0.48; I^2^= 20%; for dominant model OR_GG+AGvs.AA_= 0.94; 95%CI: 0.85-1.03; p= 0.20; I^2^= 48.08%; and for recessive model OR_AA+AGvs.GG_= 1.21; 95%CI= 0.91-1.61; p= 0.18; I^2^= 73.74%) (Figure 2, Table 3).

**Table 3:** Summary estimates for the odds ratio (OR) of *MTRR* A66G in various allele/genotype contrasts, the significance level (p value) of heterogeneity test (Q test), and the I^2^ metric

| | Genetic Contrast | Fixed effect<br>OR (95% CI), p | Random effect<br>OR (95% CI), p | Heterogeneity<br>p-value (Q<br>test) | $I^2$ (%) | Publication<br>Bias (p of<br>Egger's test) |
| --- | --- | --- | --- | --- | --- | --- |
| **All** | Allele Contrast (G vs. A) | 0.99 (0.93-1.05), 0.81 | 1.03 (0.90-1.18), 0.59 | 0.009 | 62.43 | 0.40 |
|  | Dominant (GG+AG vs. AA) | 0.94 (0.85-1.03), 0.20 | 0.94 (0.79-1.12), 0.51 | 0.06 | 48.08 | 0.42 |
|  | Homozygote (GG vs. AA) | 0.95 (0.83-1.08), 0.48 | 0.97 (0.81-1.17), 0.78 | 0.26 | 20 | 0.95 |
|  | Co-dominant (AG vs. AA) | 0.93 (0.84-1.02), 0.15 | 0.90 (0.74-1.10), 0.32 | 0.02 | 56.11 | 0.22 |
|  | Recessive (AA+AG vs. GG) | 1.06 (0.95-1.18), 0.28 | 1.21 (0.91-1.61), 0.18 | <0.001 | 73.74 | 0.32 |
| **Asian** | Allele Contrast (G vs. A) | 1.23 (1.01-1.50), 0.03 | 1.24 (0.81-1.89), 0.31 | 0.01 | 77.76 | 0.94 |
|  | Dominant (GG+AG vs. AA) | 0.99 (0.74-1.33), 0.98 | 0.99 (0.74-1.33), 0.98 | 0.68 | 0 | 0.32 |
|  | Homozygote (GG vs. AA) | 1.12 (0.70-1.81), 0.62 | 1.13 (0.70-1.81), 0.61 | 0.60 | 0 | 0.13 |
|  | Co-dominant (AG vs. AA) | 0.93 (0.68-1.26), 0.64 | 0.86 (0.55-1.33), 0.51 | 0.19 | 38.95 | 0.08 |
|  | Recessive (AA+AG vs. GG) | 1.93 (1.36-2.72), <0.001 | 1.58 (0.65-3.84), 0.30 | 0.003 | 82.48 | 0.22 |
| **Caucasian** | Allele Contrast (G vs. A) | 0.97 (0.90-1.03), 0.39 | 0.97 (0.90-1.03), 0.39 | 0.50 | 0 | 0.67 |
|  | Dominant (GG+AG vs. AA) | 0.94 (0.85-1.05), 0.32 | 0.94 (0.85-1.05), 0.31 | 0.72 | 0 | 0.21 |
|  | Homozygote (GG vs. AA) | 0.94 (0.82-1.09), 0.45 | 0.94 (0.82-1.09), 0.45 | 0.46 | 0 | 0.70 |
|  | Co-dominant (AG vs. AA) | 0.95 (0.85-1.06), 0.35 | 0.95 (0.85-1.06), 0.35 | 0.75 | 0 | 0.18 |
|  | Recessive (AA+AG vs. GG) | 0.97 (0.86-1.10), 0.72 | 0.98 (0.86-1.11), 0.75 | 0.35 | 3.89 | 0.94 |
| **Mixed** | Allele Contrast (G vs. A) | 0.93 (0.72-1.18), 0.56 | 0.91 (0.59-1.38), 0.65 | 0.09 | 64.81 | NA |
|  | Dominant (GG+AG vs. AA) | 0.78 (0.55-1.12), 0.19 | 0.42 (0.05-3.11), 0.39 | <0.001 | 91.48 | NA |
|  | Homozygote (GG vs. AA) | 0.81 (0.42-1.55), 0.53 | 0.56 (0.08-3.57), 0.54 | 0.01 | 82.09 | NA |
|  | Co-dominant (AG vs. AA) | 0.73 (0.50-1.07), 0.11 | 0.40 (0.05-2.99), 0.37 | <0.001 | 91.33 | NA |
|  | Recessive (AA+AG vs. GG) | 1.25 (0.70-2.24), 0.44 | 1.25 (0.70-2.24), 0.44 | 0.86 | 0 | NA |

**Figure 2.**
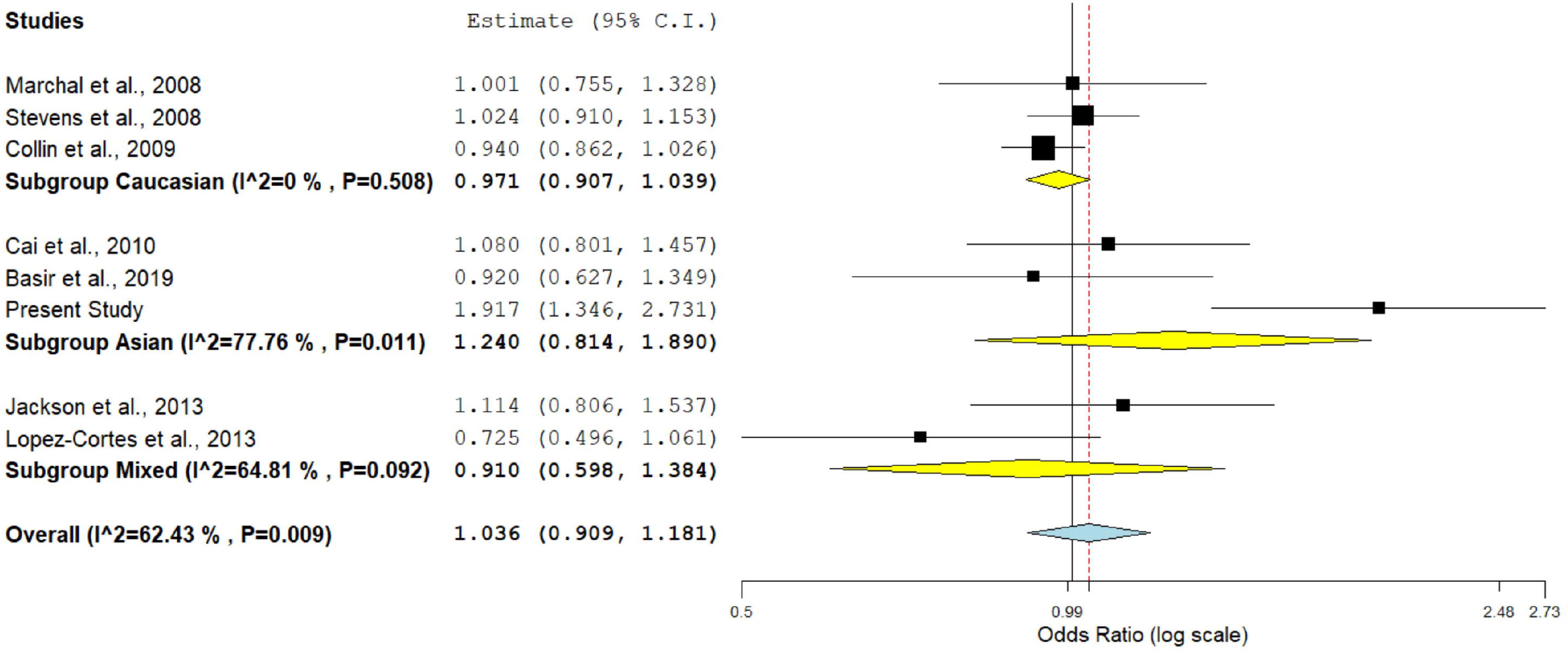
Random effect forest plot of allele contrast model (G vs. A) of *MTRR* A66G gene polymorphism.

n sub-group meta-analyses, no statistically significant association were observed in the Asian population (for G vs. A: OR= 1.24; 95% CI= 0.81-1.89; p= 0.31; I^2^= 77.76%; for GG+AG vs. AA: OR= 0.99; 95% CI= 0.74-1.33; p= 0.98; I^2^= 0%; for GG vs. AA: OR= 1.12; 95% CI= 0.70-1.81; p= 0.62; I^2^= 0%; for AG vs. AA: OR= 0.93; 95% CI= 0.68-1.26; p= 0.64; I^2^= 38.95%; and for AG+AA vs. GG: OR= 1.58; 95% CI= 0.65-3.84; p= 0.30; I^2^= 82.48%) and in the Caucasian population (for G vs. A: OR= 0.97; 95% CI= 0.90-1.03; p= 0.39; I^2^= 0%; for GG+AG vs. AA: OR= 0.94; 95% CI= 0.85-1.05; p= 0.32; I^2^= 0%; for GG vs. AA: OR= 0.94; 95% CI= 0.82-1.09; p= 0.45; I^2^= 0%; for AG vs. AA: OR= 0.95; 95% CI= 0.85-1.06; p= 0.35; I^2^= 0%; and for AG+AA vs. GG: OR= 0.97; 95% CI= 0.86-1.10; p= 0.72; I^2^= 3.89%) (Table 3).

### 3.4 Sensitivity analysis

Those studies which were not in the agreement with Hardy-Weinberg equilibrium (HWE) were excluded to conduct the sensitivity analysis. For *MTHFR* C677T gene polymorphism six studies were not in HWE [39, 42, 44, 45, 61, 65]. After removal of these studies no significant association was found in any genetic model in overall and in sub-group analyses. Similarly for the *MTRR* A66G gene polymorphism two studies were deviated from HWE [45, present study]. We removed both the studies and found no significant association in overall as well as in any sub-group meta-analyses.

### 3.5 Publican bias

Symmetrical funnel plots were observed in all five contrast genetic models (Figure 3). The p-values of Egger’s test were more than 0.05 in all the genetic contrast models which statistically confirms the symmetry of funnel plots in the overall and sub-group meta-analyses (Table 3).

**Figure 3.**
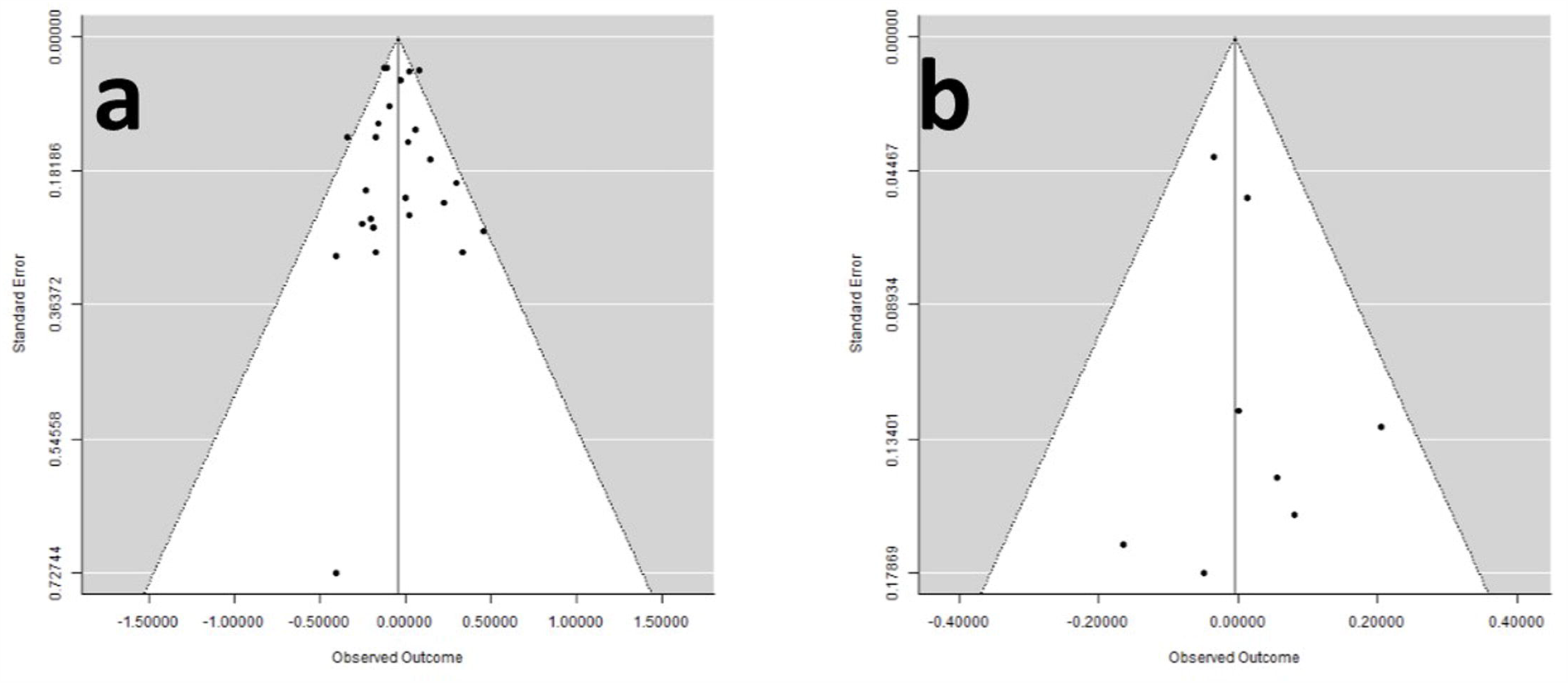
Funnel plots of standard error by log odds ratio- a) for *MTHFR* C677T gene polymorphism; b) for *MTRR* A66G gene polymorphism.

## 4. Discussion

The concentration of folate is largely determined by dietary intakes [68], and low circulating level of folate increases the risk of colon, breast and pancreatic cancer [69], while the risk of other cancers may be increases in a high folate concentration [70]. Folate is responsible for the synthesis, repair and methylation of DNA [49, 71-74]. Folate deficiency may leads to an increase in the mis-incorporation of uracil base in DNA. During repair of uracil in DNA, a transient nick is formed and two opposing nicks could lead to a break in the chromosome which could contribute to the increase risk of cancer [3]. These findings were further confirmed in the studies conducted on the lymphocytes of rats that had been maintained on a folate-deficient diet [75, 76]. Various enzymes of the folate pathway are coded by the different genes and polymorphisms in those genes may lead to differential activity of the enzymes. The most studied gene polymorphisms are *MTHFR* C677T and *MTRR* A66G.

The results of the present case-control study shows that the CT genotype (OR= 1.92; 95% CI: 1.06-3.48, p= 0.02) and the dominant model (TT+CT) (OR= 1.85; 95% CI: 1.05-3.30, p= 0.03) of the *MTHFR* C677T gene polymorphism were significantly associated with the etiology of prostate cancer. While the T allele of the *MTHFR* C677T polymorphism was marginally associated with the prostate cancer (OR= 1.67; 95% CI: 0.99-2.84, p= 0.05). Our results supported the findings of various previous studies [36, 38, 39, 41, 64].

In case of the *MTRR* A66G gene polymorphism the G allele was found to be strongly associated with the etiology of the prostate cancer (OR= 1.92; 95% CI: 1.35-2.73, p= 0.0002). Our result also confirmed the outcome of various previous published studies [39, 40, 45, 48, 49, 62].

The results of present meta-analysis showed no evidence of association of *MTHFR* C677T and *MTRR* A66G gene polymorphisms in overall and sub-group analyses with prostate cancer susceptibility.

During literature search, we found seven meta-analyses [48, 77-82] that were examined the effect of *MTHFR* C677T gene polymorphism in prostate cancer risk, but no consistent conclusion was achieved. Except two studies [80, 81], other five meta-analyses [48, 77-79, 82] reported no significant association between *MTHFR* C677T polymorphism and prostate cancer risk. Abedinzadeh et al. [80] have found significant association only in the Asian population (OR= 1.299; 95% CI= 1.121-1.506; p= 0.001). Chen et al. [81] have reported significant association between *MTHFR* C677T polymorphism with prostate cancer risk in the East Asian population using co-dominant model (CT vs. CC+TT: OR= 1.32; 95%CI= 1.02-1.70; p= 0.03). For *MTRR* A66G gene polymorphism, one meta-analysis was found during the literature search [48]. They conducted their meta-analysis with four studies and found no significant association in overall analysis.

Meta-analysis is a tool which combines different small clinical trials and increases the power of the study by reducing the type I and II errors. In recent time, meta-analysis becomes the favorite choice of researchers and numerous meta-analyses were published in past decades e.g. Down syndrome [83, 84], neural tube defects [85], Glucose 6-phosphate dehydrogenase deficiency [86], osteoporosis [87], bipolar disorder [88], depression [89], schizophrenia [90, 91], Alzheimer [92], epilepsy [93], breast cancer [94, 95], colorectal cancer [96], esophageal cancer [97], and prostate cancer [98].

The present meta-analysis has some merits over the other previously published meta-analyses such as-i) this is the largest study on the bases of number of included studies (25 studies for *MTHFR* C677T and eight studies for *MTRR* A66G) as well as on the bases of number of subjects included (26,394 subjects for *MTHFR* C677T and 8,707 subjects for *MTRR* A66G); ii) two gene polymorphisms (*MTHFR* C677T and *MTRR* A66G) are considered in meta-analysis; iii) no publication bias was found; and iv) studies were searched by using four different databases *viz*. PubMed, Science Direct, Springer Link and Google scholar. Here we also want to acknowledge few limitations of the meta-analysis like i) we used only crude odds ratios; ii) only English language publications were included; and iii) the effect of the hyperhomocystenemia and folate deficiency were not considered.

## 5. Conclusions

The data of our case-control study revealed that the T allele, CT genotype, and dominant (TT+CT) model of *MTHFR* C677T gene polymorphism and G allele of *MTRR* A66G gene polymorphism were significantly associated with the etiology of prostate cancer risk. In future, larger case-control association studies from different global populations are required to find out the exact association between these two polymorphisms (C677T and A66G) and prostate cancer risk. In addition, effects of higher concentration of homocysteine and lower concentration of folate should also be evaluated in prostate cancer patients.

## Data Availability

All data will be provided in request to the corresponding author.

## Acknowledgments

Upendra Yadav is highly grateful to VBS Purvanchal University, Jaunpur for providing financial assistance to him in the form of PDF.

